## Supporting Information for "Genomics reveals heterogeneous *Plasmodium falciparum* transmission and population differentiation in Zambia and bordering countries"

### SI Appendix includes:

- Figures S1 to S12
- Tables S1 to S8 (provided separately as additional file in Excel format)

**Supplementary Figures**

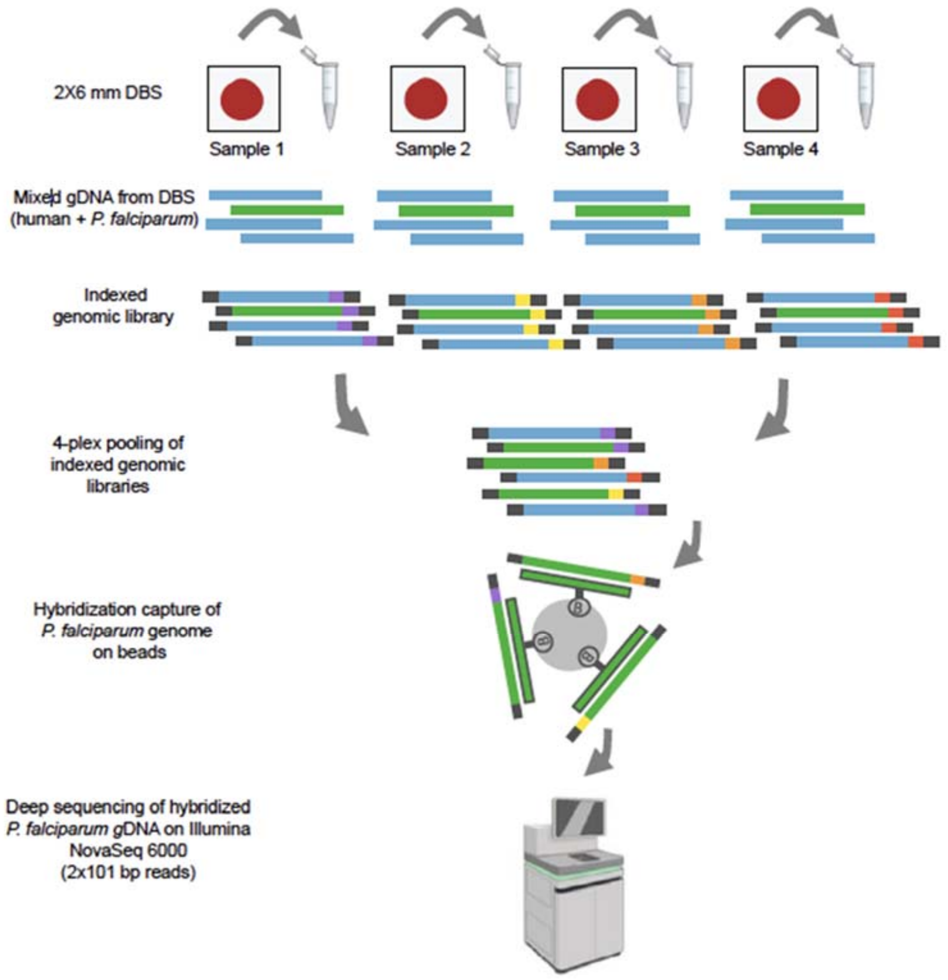


**Figure S1. Multiplexed *P. falciparum* whole-genome capture and sequencing from DBS samples.** Schematic representation of 4-plex *P. falciparum* whole-genome capture and deep sequencing. Dual-Indexed genomic libraries were created from gDNA extracts from human DBS *P. falciparum* positive samples (mixed DNA template in blue and green colors), pooled in 4-plex and subsequently incubated with custom probes (SeqCap EZ). *P. falciparum* custom probes were designed to tile 98% *P. falciparum* 3D7 reference genome to enable target enrichment of *P. falciparum* DNA without amplification biases. Above, *P. falciparum* DNA (Green) hybridized to the probes while non-targeted DNA (Blue) is washed off. Hybridized DNA is then deep sequenced on Illumina NovaSeq 6000 platform.

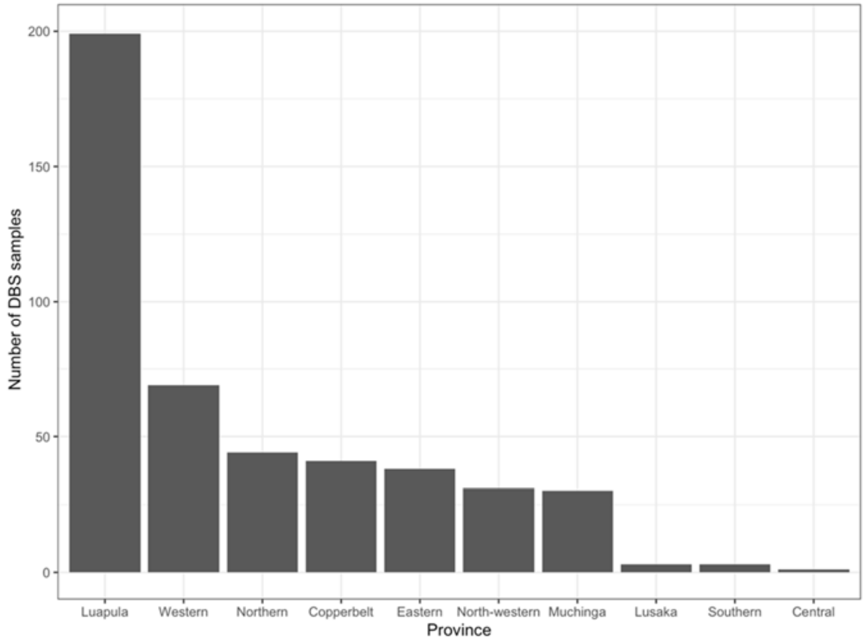


**Figure S2. Distribution of WGS sequenced *P. falciparum* samples per province across Zambia.** A total of 459 (114 capture reactions-4plex and 1 capture reaction-3-plex) samples were sequenced from 10 provinces. Number of sequenced samples between provinces varied as a function of sampling efforts (see Methods for details).

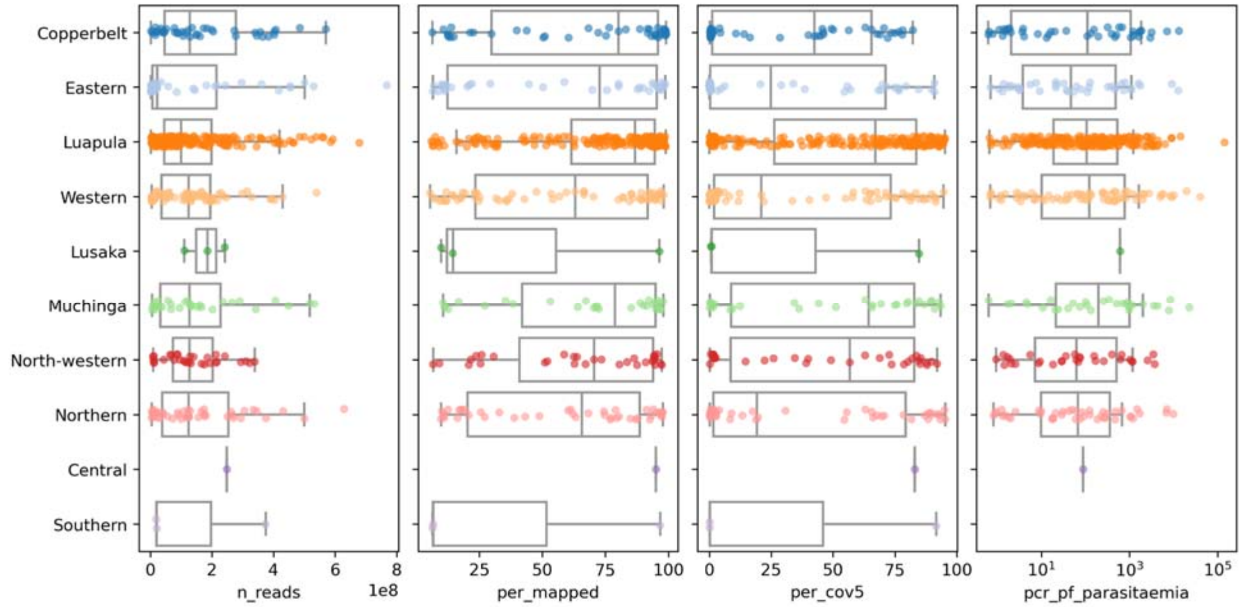


**Figure S3. Sequence metrics across 459 *P. falciparum* WGS sequenced samples by geographic origin.** From left to right, in the boxplots the x-axes represent the number of sequenced reads, the percentage of read mapping to *P. falciparum* 3D7 reference genome, the percentage of genome with minimum read depth of 5X, and the parasitemia estimated by PET-PCR (parasites/□L, log scale). The y-axis represents the geographic origin of the DBS samples at the provincial level. For each boxplot the middle line represents the median value of metrics of interest, the box represents the interquartile range, and the whiskers represent the range excluding outliers.
Five samples (2 samples out of 3 from Lusaka Province and all 3 samples from Southern Province) were malaria positive by RDT but we did not have an estimated parasitemia by PET-PCR, therefore were omitted from the last bloxplot on the right.

**A.**

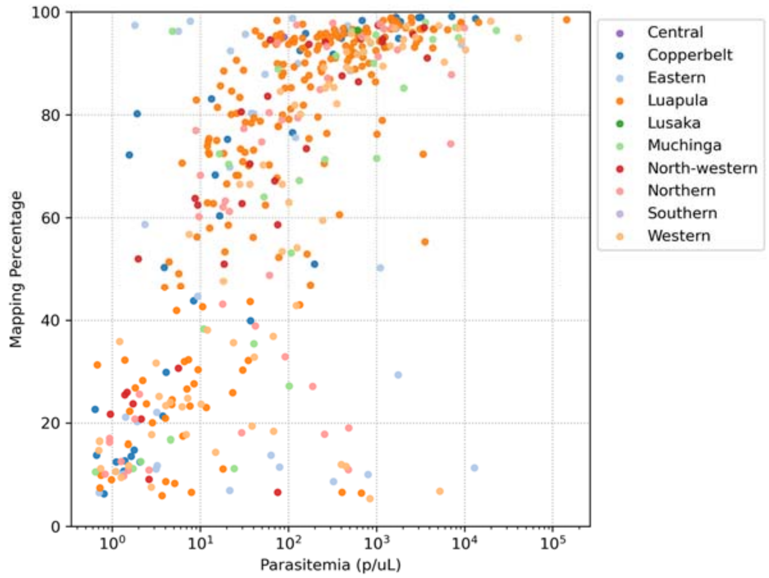


**B.**

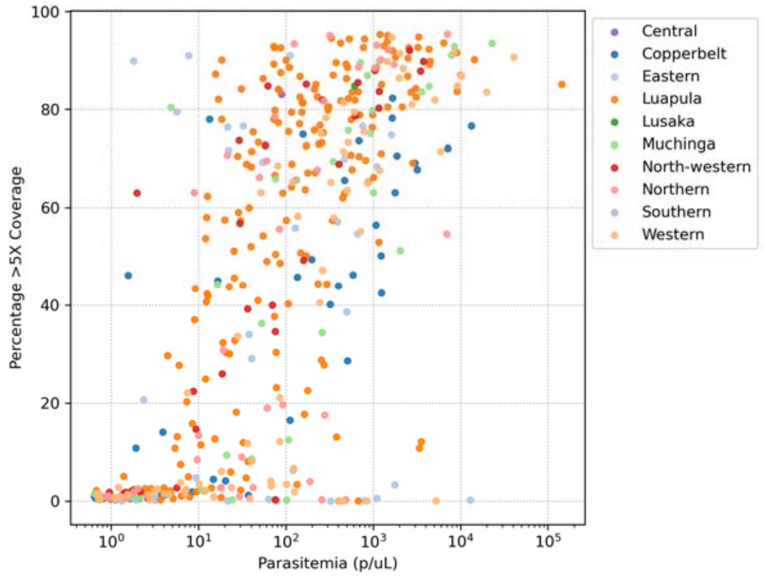


**Figure S4. Correlates of *P. falciparum* capture efficiency and genome coverage**. WGS samples are represented by dots and colors denote sample origin **(A)** *P. falciparum* capture efficiency (the percentage of read mapping to *P. falciparum* 3D7 reference genome) is plotted against the estimated parasitemia (measured by PET-PCR). **(B)** The percentage of *P. falciparum* genome coverage with a minimum of 5X read coverage is plotted against the estimated parasitemia. *P. falciparum* parasitemia is a significant predictor of capture efficiency and genome coverage in univariate quasi-Poisson models.

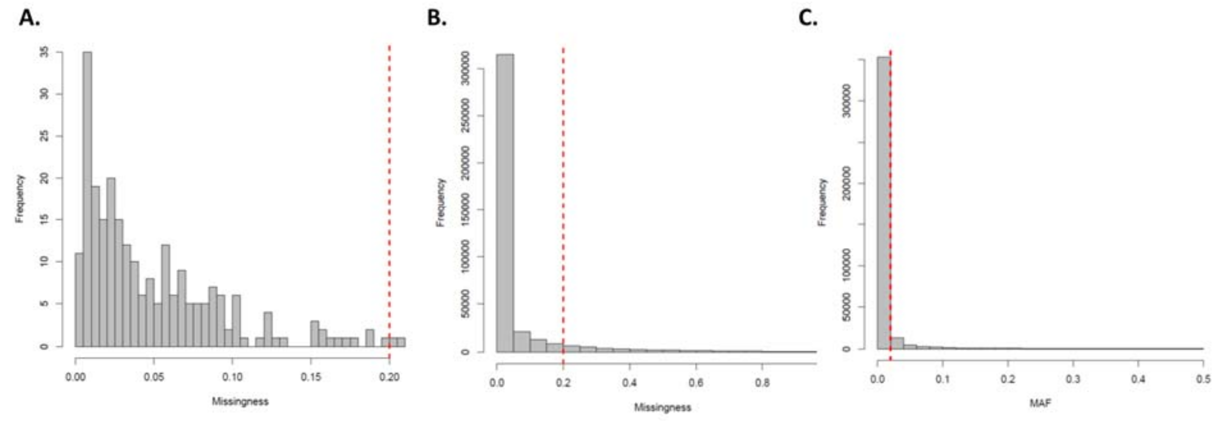


**Figure S5. Sample missingness (A), SNP missingness (B) and Minor allele frequency (C) distributions and filtering threshold**. Prior to variant filtering we scored 1,121,403 SNPs with a VQSLOD >0 across the 241 genomes. After filtering out variants in telomeric regions, a total of 389,097 biallelic SNPs were retained across the *P. falciparum* core genome, 358,260 SNPs remained after 0.2 missingness filtering and 29,992 SNPs remained after MAF 0.02 filtering.

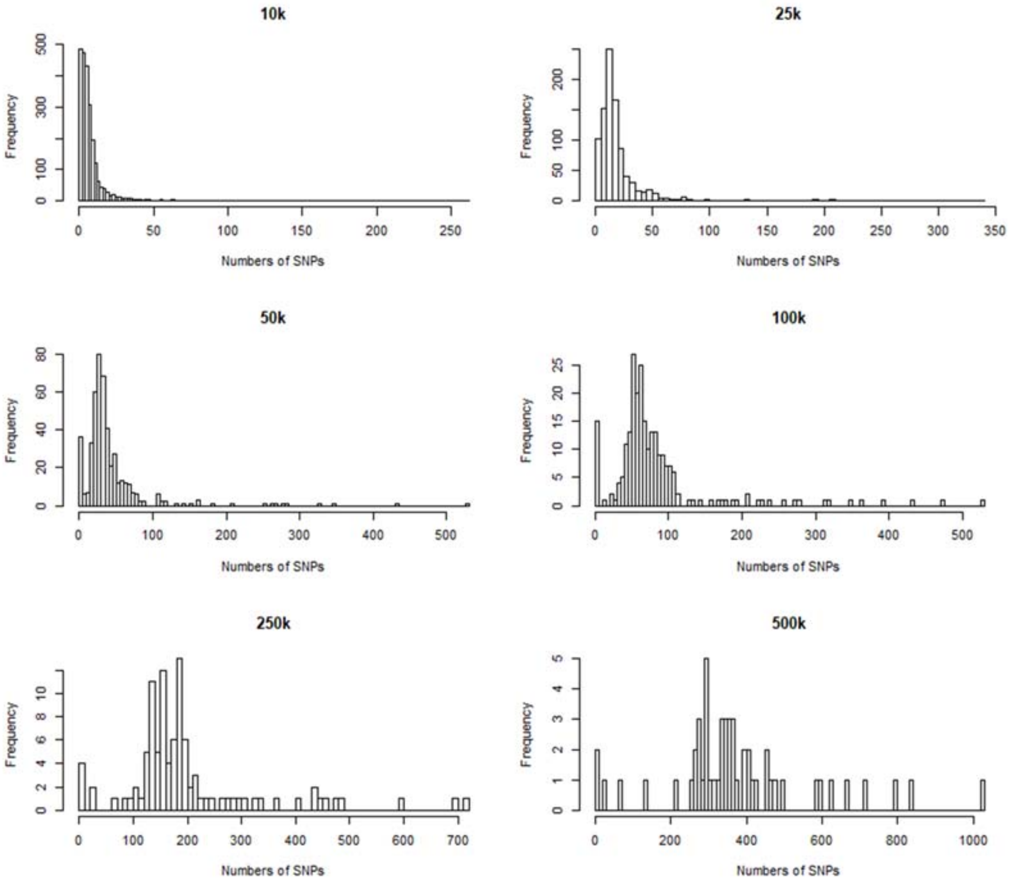


**Figure S6. Distribution of 29,992 genome-wide SNPs using different window sizes.** Each SNP frequency plot illustrates a different window size from 10K to 500Kb. The x-axis indicates number of SNPs and y-axis indicates frequency of SNPs across sequenced samples.

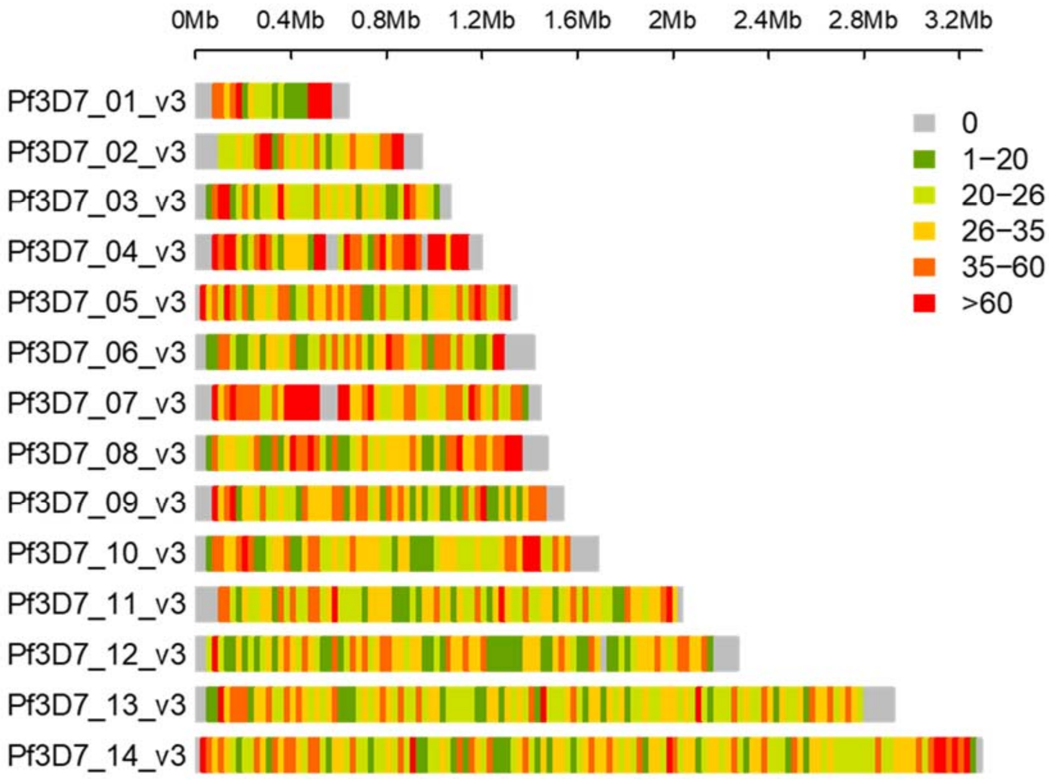


**Figure S7. Genome-wide SNP density plot.** The plot shows the SNP density plot chromosome wise representing the number of SNPs within 25Kb window size. The horizontal axis denotes the chromosome length (Mb), and the different colors depict SNP density from low (green) to high (red). Masked telomeric and hypervariable regions are shown in gray.

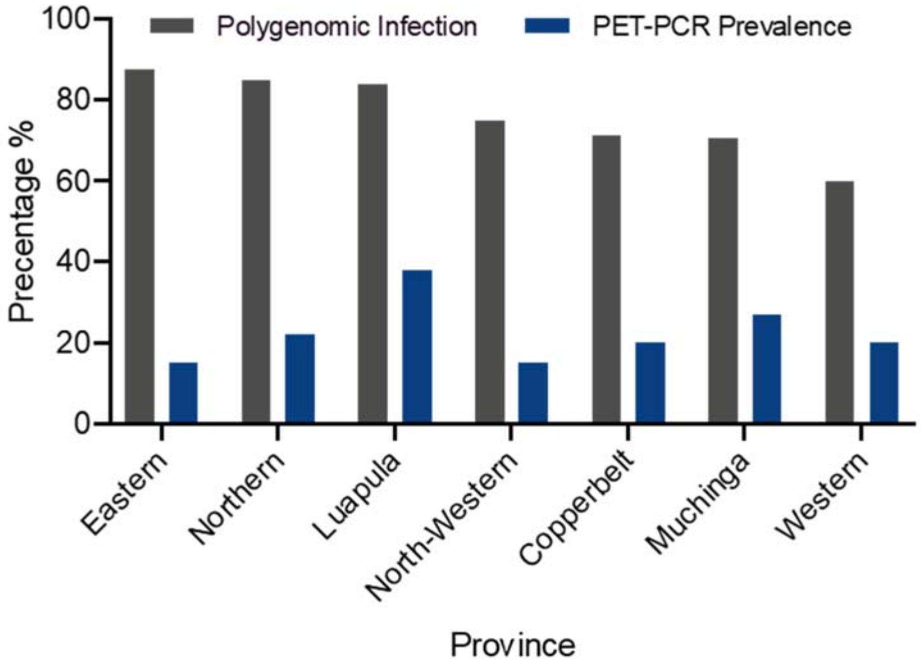


**Figure S8. *P. falciparum* malaria prevalence and polygenomic infections per province.** The x-axis indicates provinces and y-axis indicate percentage of polygenomic infections (grey bars) (number of samples harboring more than one distinct parasite genome, $F_{WS} < 0.95$, divided by total number of samples sequenced per province X100). Blue bars represent *P. falciparum* malaria prevalence as estimated by PET-PCR.

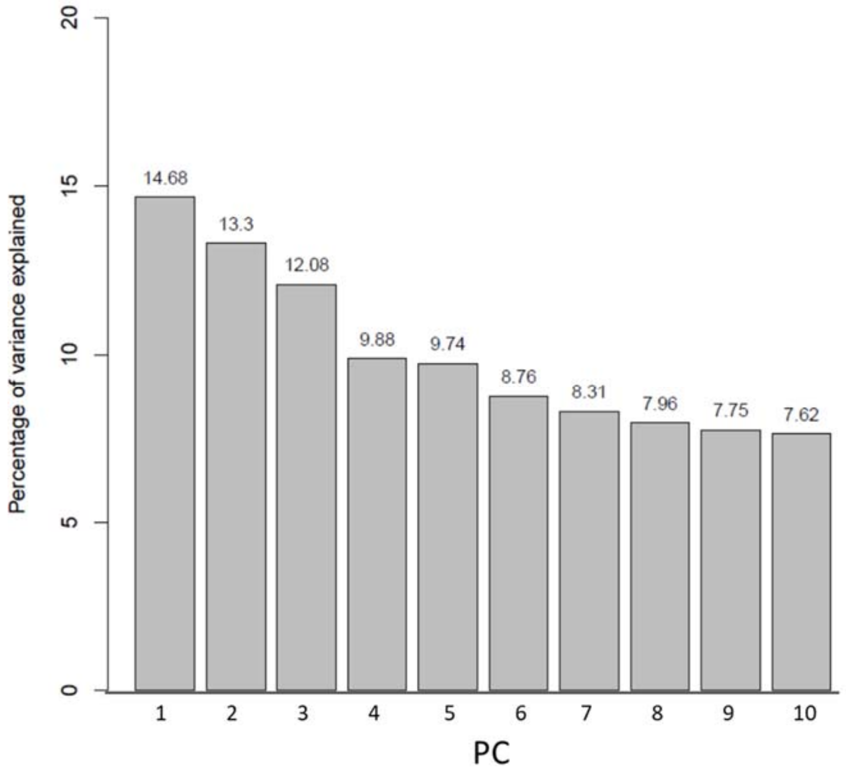


**Figure S9. Percentage of variance explained by the Principal Components (PC) for Zambian *P. falciparum* parasites**. Bar plot shows the first 10 PCs and the percentage of variance explained by each.

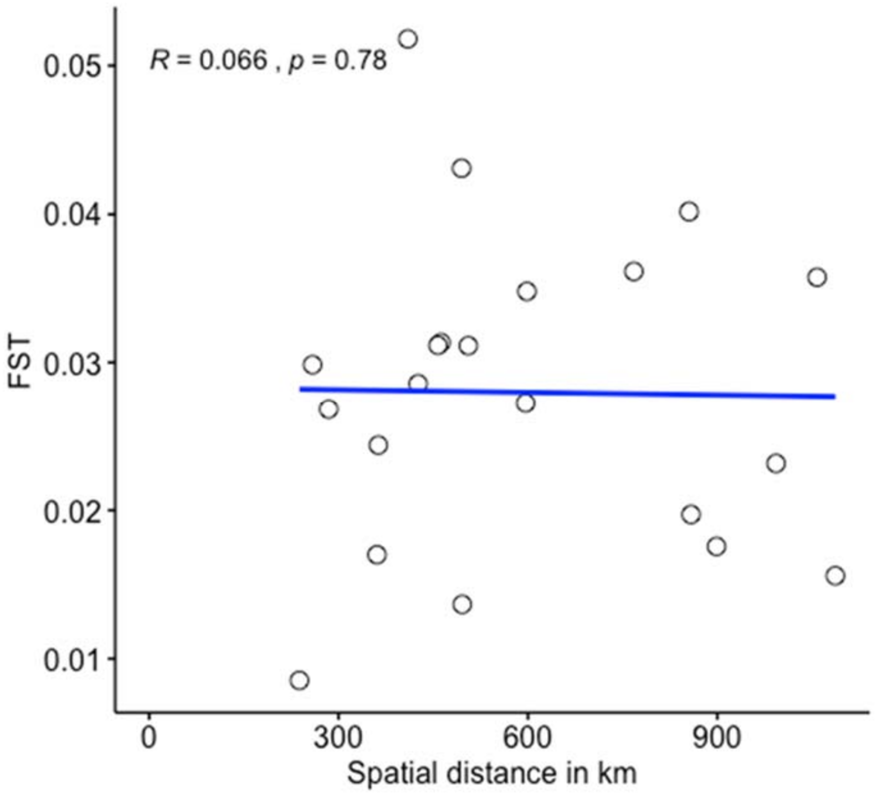


**Figure S10. Mantel test showing the relationship between genetic distance and geographical distance.** Pairwise $F_{ST}$ value (genetic differentiation) and actual geographic distance between seven provinces across Zambia used for this analysis.

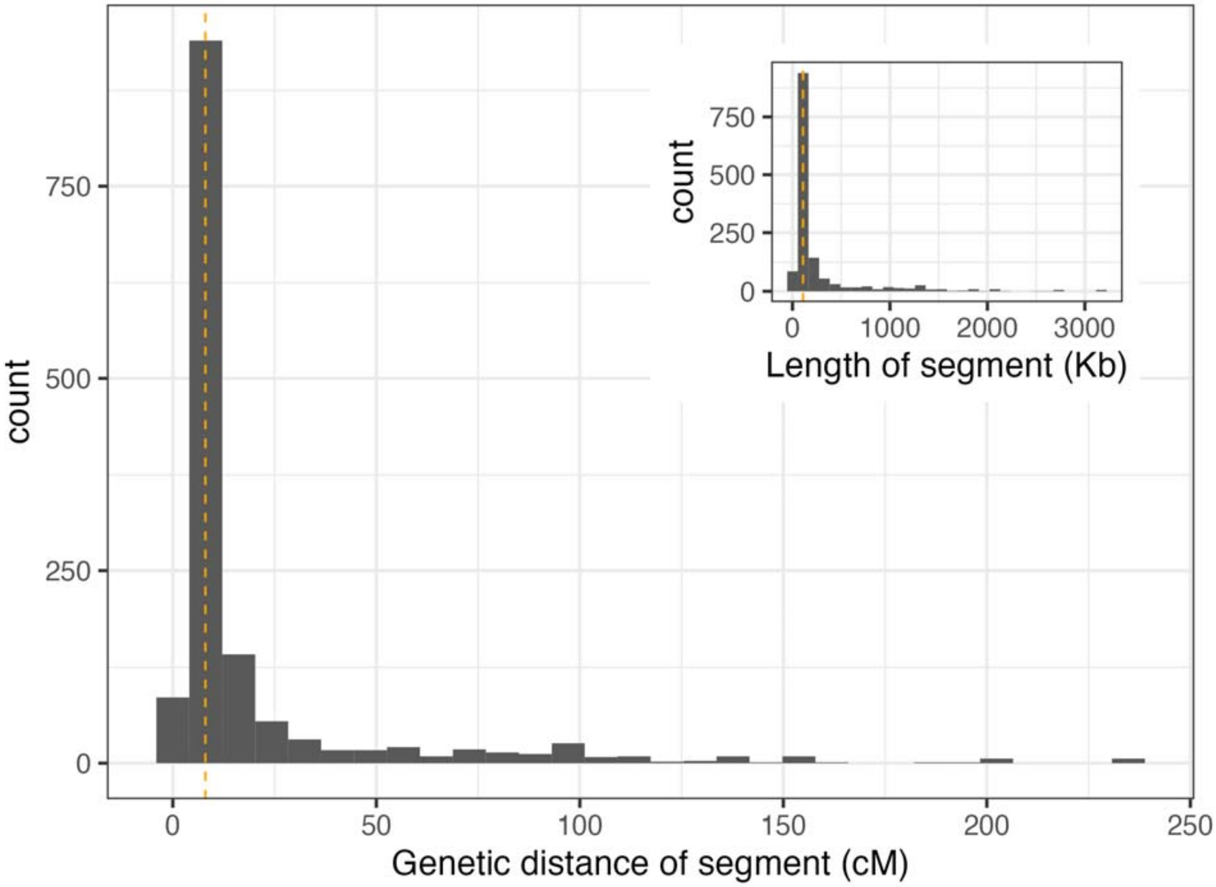


**Figure S11. Genetic distance and length (inset) of pairwise IBD segments across the 241 *P. falciparum* sequenced genomes.** The genetic distances (measured in centimorgan [cM] are calculated by assuming a constant recombination rate across the *P. falciparum* genome (13.5Kb/cM). The x-axis represents the length of IBD genomic segments in cent Morgan (cM) and the y-axis represent their frequency across all samples. The vertical dashed red line represents the median genetic distance, equivalent to 8 cM, which corresponds to approximately to six generations (range= 3-239cM).

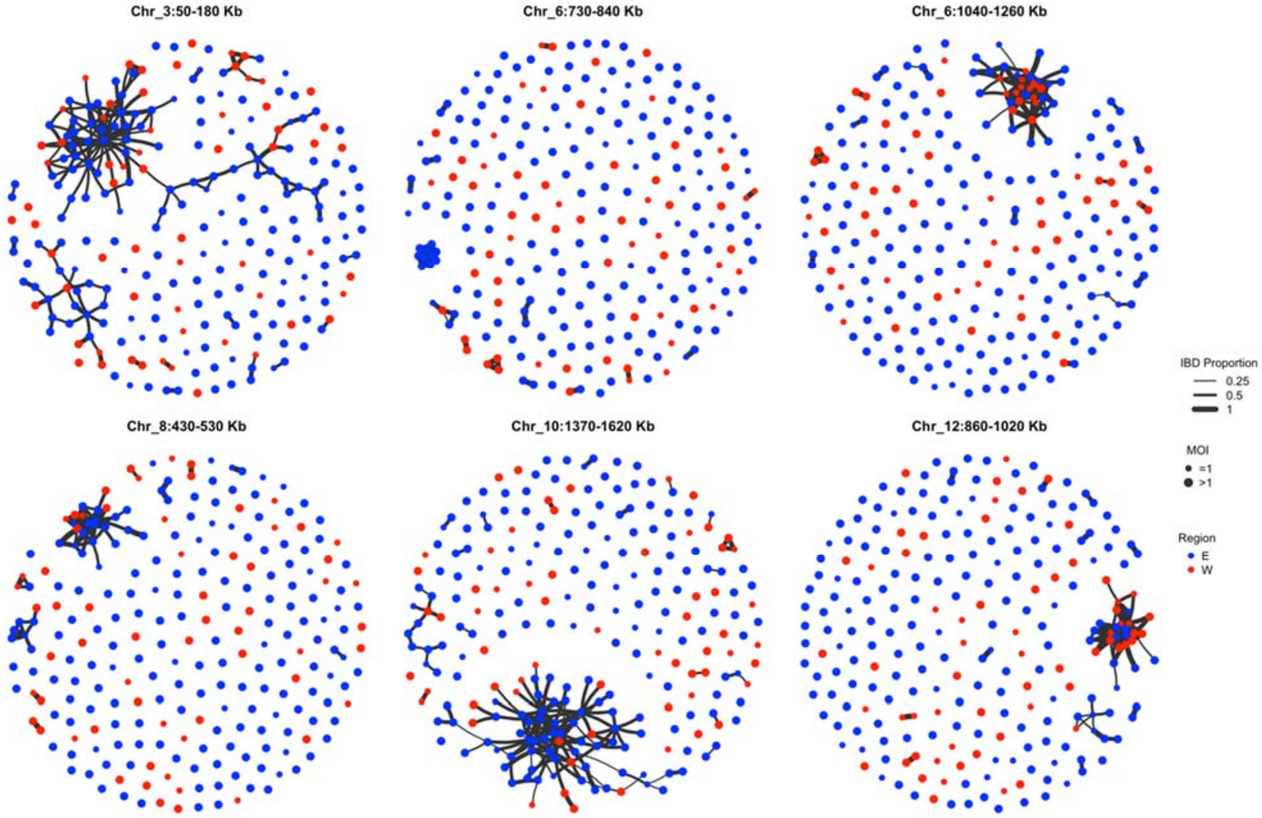


**Figure S12. IBD pattern among samples in regions identified under selection using $X_{iR}$ statistics**. Proportion of IBD sharing is calculated for the specified chromosomal regions above each subgraph. Only edges with IBD sharing >= 25% are included. Blue dots represent samples from the Eastern provinces (Luapula, Northern, Muchinga, Eastern). Red dots represent samples from the Western provinces (North-Western, Copperbelt, Western, Central, Lusaka, Southern). Size of the dots represents the mixed infectious status (MOI) of the sample.

### Supplementary Tables

**Table S1. Sample description.** The table describes 241 *P. falciparum* WGS sample successfully sequenced (defined as >50% of *P. falciparum* genome with > 5X read coverage) from Zambia, sample origin (province, district and ward levels), cluster ID (smallest geographical unit used in the MIS 2018), cluster coordinates, $F_{WS}$ value and their polygenomic status (i.e. $F_{WS}$ < 0.95 classified as polygenomic and $F_{WS}$ >0.95 classified as monogenomic).

**Table S2. Polygenomic infection and parasite prevalence data at the cluster level across Zambia.** The table describes the number of *P. falciparum* WGS samples retained from Zambia per geographic cluster (smallest geographic unit according to the MIS 2018 data collection). Percentage of polygenomic infections (number of samples harboring more than one distinct parasite genome ($F_{WS}$ <0.95) divided by total number of samples sequenced per province X100).

**Table S3. Polygenomic infection and parasite prevalence data at the provincial level.** The table describes the number of *P. falciparum* WGS samples retained from Zambia per province (Central, Lusaka and Sonuthern provinces are excluded due to the small sample size, see Methods). Percentage of polygenomic infections (number of samples harboring more than one distinct parasite ($F_{WS}$ <0.95) divided by total number of samples sequenced per province X100).

**Table S4. Pairwise population differentiation among *P. falciparum* populations across seven provinces in Zambia**. Numbers in the upper right indicate pairwise genetic differentiation ($F_{ST}$) values calculated on the genome-wide SNP data set.

**Table S5. List of the significant selected SNPs across six regions in the chromosomes 3, 6, 8 10 and 12 of the *P. falciparum* genome.** This table contains the significant SNPs (snp_id), location of the SNP, XiR, P value and -log10 transformed P-value, and their annotations (original EFFtag, function, Codon changes, amino acid change, gene names, exon number, etc.)

**Table S6. Range info of significant regions.** This table contains the coordinates and SNP statistics of the *P. falciparum* genomic regions with at least two significantly selected SNPs within a window size of 50Kb.

**Table S7. List of *P. falciparum* genome samples downloaded from the Pf3k database and included for the continental analysis in this study.** Sample ID, metadata and sequencing metrics for 714 *P. falciparum* genomes from four African countries (Democratic Republic of Congo, Ghana, Guinea, and Malawi) included in the continental analysis are reported.

**Table S8. List of *P. falciparum* genome samples downloaded from SRA originated from Tanzania and included for the continental analysis in this study.** Sample ID, metadata and sequencing metrics for 68 *P. falciparum* genomes from Tanzania included in the continental analysis are reported.

**Table S9. List of inferred CNV for significantly selected genes calculated from monogenomic samples.** This table contains chromosome, gene location, gene ID, gene length, gene name, gene product description, mean CNV, median CNV, standard deviation of CNV and coefficient of variation of CNV.
